# Multiple breath-washout for pulmonary function assessment in young childhood cancer survivors: a multicenter study

**DOI:** 10.1101/2025.11.17.25340423

**Authors:** Maša Žarković, Christina Schindera, Nicolas Waespe, Jakob Usemann, Christine Schneider, Anne Mornand, Marc Ansari, Philipp Latzin, Claudia E Kuehni

**Affiliations:** Childhood Cancer Research Group, Institute of Social and Preventive Medicine, University of Bern, Switzerland; Graduate School for Health Sciences, University of Bern, Switzerland; Division of Paediatric Oncology/Haematology, University Children’s Hospital Basel, University of Basel, Switzerland; Division of Pediatric Hematology and Oncology, Department of Pediatrics, Inselspital, Bern University Hospital, University of Bern, Switzerland; CANSEARCH Research Platform for Pediatric Oncology and Hematology, Department of Pediatrics, Gynecology and Obstetrics, Faculty of Medicine, University of Geneva, Switzerland; Division of Pulmonology, University Children’s Hospital Basel (UKBB), University of Basel, Switzerland; Department of Respiratory Medicine, University Children’s Hospital Zurich, University of Zurich, Switzerland; Children’s Research Center, University Children’s Hospital Zurich, University of Zurich, Switzerland; Division of Pediatric Pulmonology, Department of Women, Child and Adolescent, University Hospital of Geneva, Switzerland; Division of Pediatric Oncology and Hematology, Department of Women, Child and Adolescent, University Hospital of Geneva, Switzerland; Division of Paediatric Respiratory Medicine and Allergology, Department of Paediatrics, Inselspital, University Hospital, University of Bern, Switzerland

## Abstract

**Rationale:** Childhood cancer survivors (CCS) are at risk for long-term pulmonary complications from treatment-related toxicity. Nitrogen multiple-breath washout (N_2_MBW) may detect small airway dysfunction earlier than standard pulmonary function tests (PFTs), but its value in CCS remains uncertain.

**Objectives:** To determine the prevalence of ventilation inhomogeneity measured by N_2_MBW in CCS, evaluate whether it detects abnormalities beyond spirometry and diffusion capacity for carbon monoxide (DLCO), and explore associations with treatment exposures.

**Methods:** In this prospective multicenter study (Bern, Basel, Geneva), we measured lung function in CCS aged 6–21 years who were undergoing routine follow-up. We stratified participants into high-risk (pulmotoxic chemotherapy [busulfan, bleomycin, nitrosoureas], thoracic surgery or radiotherapy, or hematopoietic stem cell transplantation [HSCT]) and standard-risk (other systemic anticancer treatments). PFTs included N_2_MBW (lung clearance index [LCI], acinar [S_ACIN_] and conductive [S_COND_] inhomogeneity), spirometry (forced expiratory volume in 1 second [FEV₁], forced vital capacity [FVC]), and DLCO. Abnormal values were defined as z-score <-1.645 or >1.645, calculated using the Global Lung Initiative references. We quantified the proportion of participants with isolated elevated LCI and analyzed associations with treatment exposures using multivariable linear regression.

**Results:** We included 191 CCS (median 7 years post-diagnosis). Mean LCI was 6.27 (95%CI 6.17–6.37), with 7% abnormal. N_2_MBW results did not differ between high- and standard-risk groups, but allogeneic HSCT survivors had the highest mean LCI (7.18, 95%CI 5.88–8.37), with 33% abnormal. Survivors had mildly impaired mean FEV_1_, FVC, and DLCO z-scores (−0.21, −0.34, 0.23), more pronounced in high-risk CCS (−0.67, −0.85, −0.05). Isolated LCI impairment, without abnormalities in spirometry or DLCO, occurred in only 3%. Among treatment exposures, only allogeneic HSCT was associated with higher LCI (1.40, 95%CI 0.57–2.33) and S_ACIN_ (1.24, 95%CI 0.20–2.28).

**Conclusion:** Most pediatric CCS had normal lung function. N_2_MBW abnormalities were rare, occurring mainly in allogeneic HSCT survivors. Overall, N_2_MBW added little beyond standard PFTs, suggesting it may not be needed for routine follow-up at this stage, except after allogeneic HSCT. Larger, longitudinal studies should clarify the onset, progression, and prognostic significance of N_2_MBW abnormalities and their potential role in risk-adapted follow-up care.

## Introduction

Pulmonary late effects are a significant cause of long-term morbidity and reduced quality of life in childhood cancer survivors (CCS) (1). Several cancer treatments, including chemotherapies such as bleomycin, busulfan, and nitrosoureas, thoracic radiotherapy or surgery, and hematopoietic stem cell transplantation (HSCT), damage different anatomical compartments of the lung (2,3). These treatments can injure the alveolar epithelium, pulmonary vasculature, and interstitial tissue through a range of mechanisms, including oxidative stress, inflammation, and fibrosis (4,5), resulting in permanent structural changes and progressive functional impairment (6,7). To monitor pulmonary health in this at-risk population, current surveillance guidelines from the Children’s Oncology Group recommend spirometry and measurement of the diffusing capacity of the lungs for carbon monoxide (DLCO) (3). While spirometry primarily detects changes in large airway function (obstruction, restriction), DLCO measures gas exchange at the alveolar-capillary membrane. As such, they may miss early or subtle abnormalities in the small airways, a compartment where pathological changes may begin and remain clinically silent for years (8,9).

Nitrogen multiple-breath washout (N_2_MBW) has emerged as a promising, non-invasive technique to assess ventilation inhomogeneity, with particular sensitivity to small airway dysfunction, as shown in studies of children with cystic fibrosis or asthma (10,11). In a systematic review, we found only three studies of N_2_MBW in CCS; all were single-center, mostly retrospective, and had small sample sizes, limiting conclusions about its diagnostic value (12). In a previous study of adult long-term CCS, we found that N_2_MBW identified more abnormalities than spirometry, including among survivors not exposed to recognized lung-damaging treatments (13), suggesting that small airway disease may often go undetected by standard tests. Whether N_2_MBW is equally sensitive in younger survivors, and how it correlates with other lung function tests, remains unclear. Therefore, this study aimed to assess the prevalence of pulmonary impairment detected by N_2_MBW in a comprehensive cohort of young CCS, to evaluate whether it detects abnormalities beyond spirometry and DLCO, and to investigate associations with treatment exposures.

## Methods

### Study design and study population

We assessed data from the ongoing Swiss Childhood Cancer Survivor Study FollowUp–Pulmo (SCCSS-FU Pulmo), which investigates long-term pulmonary outcomes in CCS. The study is embedded in routine pediatric oncology follow-up care at the University Children’s Hospitals of Bern, Basel, and Geneva, Switzerland. A research nurse screens upcoming oncology follow-up appointments to identify eligible survivors, and the clinics coordinate a comprehensive pulmonary evaluation during survivors’ scheduled visits. Participants complete standardized pulmonary function tests (PFTs), including N_2_MBW. Detailed study procedures have been published (14). The present analysis focuses on the subgroup that completed N_2_MBW. Results from other PFTs in the complete SCCSS-FU Pulmo cohort have been reported (15).

Eligible for participation were CCS aged 6 to 21 years, who had been diagnosed with cancer as classified by the International Classification of Childhood Cancer, Third Edition (ICCC-3) (16), completed cancer treatment, were in remission, and attended follow-up care between June 2022 and July 2025. We excluded survivors treated solely with surgery or radiotherapy not involving the thorax, those with a current relapse, or in palliative care, and those unable to perform PFTs due to physical or cognitive limitations.

We stratified participants into two risk groups based on cancer treatments. The high-risk group included CCS who had received pulmotoxic chemotherapy (busulfan, bleomycin, carmustine, or lomustine) (2,3); thoracic surgery involving the chest or lungs (3); pulmotoxic radiotherapy (radiation to the chest (mantle, mediastinal, or whole lung fields), abdomen (whole or any upper field), or total body irradiation) (2,3); or HSCT (17). The standard-risk group included CCS treated with other systemic anticancer therapies (chemotherapy, immunotherapy, or targeted agents) (18). Additionally, we subdivided the high-risk group into HSCT survivors (autologous and allogeneic) and others, based on previous evidence suggesting that N_2_MBW may be particularly sensitive in detecting pulmonary impairment in this population (12).

All participants or their legal guardians provided written informed consent, and the study was approved by the Ethics Committee of the Canton of Bern (KEK-BE: 2019-00739).

### Pulmonary function tests

#### Nitrogen multiple breath washout test

Trained lung function technicians in all centers performed N_2_MBW measurements using the Exhalyzer D device (EcoMedics AG). Data were analyzed using Spiroware Software version 3.3.1 (EcoMedics AG). The primary outcome was the lung clearance index (LCI), which represents the number of functional residual capacity (FRC) turnovers needed to clear the lungs of nitrogen to 1/40th of the initial concentration. Secondary outcomes included ventilation heterogeneity in the convection-dependent airways (S_COND_) and the diffusion–convection–dependent airways (S_ACIN_). To account for lung size, we adjusted S_COND_ and S_ACIN_ values by multiplying them by tidal volume (V_T_). All N_2_MBW measurements underwent centralized quality control using a specialized software developed at the University of Bern (19). The software automatically detects technical artefacts such as leaks, insufficient waiting time between tests, early termination of tests, synchronization issues, and abnormal breathing patterns or volumes, in line with current consensus guidelines (10,20). We calculated z-scores for LCI using the most recent Global Lung Initiative (GLI) reference equations (21) and interpreted S_ACIN_ and S_COND_ using reference values derived from a healthy adult population (22).

#### Spirometry and DLCO

Participants completed spirometry and DLCO following N_2_MBW and according to the European Respiratory Society and American Thoracic Society (ERS/ATS) consensus statement (23). Experienced lung function technicians assessed the quality of each test. Primary outcome measures were forced expiratory volume in 1 second (FEV_1_), forced vital capacity (FVC), the Tiffenau index (FEV_1_/FVC), and forced expiratory flow at 25% to 75% of FVC (FEF_25-75%_). The main outcome for gas transfer was DLCO. We calculated z-scores using the GLI reference equations (24–26).

#### Clinical data

We extracted demographic, clinical, and treatment-related information from medical records. Anthropometric measures (height, weight, and body mass index) were assessed at the time of the PFTs and converted into z-scores using World Health Organization reference standards (27). We defined current asthma based on a documented diagnosis in medical records.

#### Statistical analysis

We described continuous variables using means with 95% confidence intervals (CI) or medians with interquartile range (IQR), and categorical variables using frequencies and percentages. Lower and upper limits of normal (LLN and ULN) were defined as z-scores <-1.645 and >1.645, respectively. To compare risk groups, we applied two-sample t-tests or Mann-Whitney U tests for continuous variables, and chi-square or Fisher’s exact tests for categorical variables, depending on data distribution and cell counts. To examine the relationship between the main N_2_MBW outcome (LCI) and spirometry and DLCO parameters, we calculated Spearman’s rank correlation coefficients and determined the proportion of participants with isolated elevated LCI. Using multivariable linear regression models, we examined the associations between treatment exposures and N_2_MBW parameters (LCI, S_COND_ x V_T_, S_ACIN_ x V_T_). Independent variables included pulmotoxic chemotherapy, thoracic surgery, pulmotoxic radiotherapy, and HSCT (autologous and allogeneic), as these modalities have been associated with long-term pulmonary complications. All models were adjusted for age at study, sex, time since cancer diagnosis, and asthma. We used Stata version 16.1 (StataCorp LLC) and R version 4.4.2 (R Foundation for Statistical Computing).

## Results

### Study population

By July 2025, 278 survivors across the three centers met eligibility criteria for the SCCSS-FU Pulmo study, and 251 (90%) participated. Within this cohort, 31 did not perform the N_2_MBW, mainly due to logistical challenges at one center, where it required an additional appointment. Among those who completed the test, measurements from 29 participants did not meet quality criteria. This resulted in high-quality N_2_MBW data for 191 participants. (Supplementary Figure S1). Participants with valid N_2_MBW measurements were slightly older (median 14 vs. 13 years) and had completed treatment longer ago (median 7 vs. 5 years since diagnosis) compared to those without N_2_MBW (Supplementary Table S1).

The median age at study enrollment was 14 years (IQR 10–17) and the median time since diagnosis was 7 years (IQR 4–10); 57% were male (Table 1). The most common cancer types were leukemia (48%), lymphoma (13%), and neuroblastoma (11%). Nearly all had received chemotherapy (98%). Twenty-one participants (11%) had received pulmotoxic chemotherapy, 27 (14%) pulmotoxic radiotherapy, 16 (8%) had undergone thoracic surgery, and 25 (13%) HSCT (autologous 7%, allogeneic 6%). Asthma was diagnosed in 6% (n=11), and one participant had bronchiolitis obliterans. Overall, 28% (n=53) were categorized as high-risk and 72% (n=138) as standard-risk.

**Table 1.** Characteristics of study participants, overall and stratified as high-risk (participants exposed to pulmotoxic chemotherapy, radiotherapy, thoracic surgery, and/or HSCT) and standard-risk (participants not exposed to pulmotoxic treatments).

|  | **Total**<br>**N=191, 100%** | **High Risk**<br>**n=53, 28%** | **Standard Risk**<br>**n=138, 72%** |
| --- | --- | --- | --- |
| **Demographic characteristics** |  |  |  |
| **Sex** |  |  |  |
| Male, n (%) | 108 (57) | 28 (53) | 80 (58) |
| Female, n (%) | 83 (43) | 25 (47) | 58 (42) |
| Age at study in years, median (IQR) | 14 (11–17) | 14 (10–16) | 14 (11–17) |
| Age at diagnosis in years, median (IQR) | 5 (3–10) | 6 (3–9) | 5 (3–10) |
| Time since diagnosis in years, median (IQR) | 7 (5–10) | 7 (5–9) | 7 (5–10) |
| **Clinical characteristics** |  |  |  |
| Weight at study, z-score median (IQR) | 0.15 (-0.39 – 1.04) | -0.15 (-0.88 – 0.53) | 0.29 (-0.30 – 1.18) |
| Height at study, z-score median (IQR) | 0.13 (-0.70 – 0.68) | -0.11 (-0.84 – 0.58) | 0.20 (-0.68 – 0.71) |
| BMI at study, z-score median (IQR)<sup>a</sup> | 0.30 (-0.35 – 1.28) | -0.21 (-0.76 – 0.86) | 0.63 (-0.30 – 1.45) |
| **BMI categories, n (%)<sup>a</sup>** |  |  |  |
| Underweight | 5 (3) | 4 (8) | 1 (1) |
| Normal weight | 127 (66) | 38 (72) | 89 (65) |
| Overweight | 41 (22) | 10 (19) | 31 (22) |
| Obese | 18 (9) | 1 (2) | 17 (12) |
| **ICCC-3 cancer diagnosis, n (%)** |  |  |  |
| Leukemia | 91 (48) | 13 (25) | 78 (57) |
| Lymphoma | 24 (13) | 5 (9) | 19 (14) |
| CNS neoplasms | 9 (5) | 5 (9) | 4 (3) |
| Neuroblastoma | 20 (11) | 12 (23) | 8 (6) |
| Renal tumors | 16 (8) | 10 (19) | 6 (4) |
| Malignant bone tumors | 7 (4) | 4 (8) | 3 (2) |
| Soft tissue sarcomas | 12 (6) | 2 (4) | 10 (7) |
| Other neoplasms<sup>b</sup> | 12 (6) | 2 (4) | 10 (7) |
| History of relapse, n (%) | 15 (8) | 11 (21) | 4 (3) |
| Any chemotherapy, n (%) | 188 (98) | 50 (94) | 138 (100) |
| Pulmotoxic chemotherapy, n (%) | 21 (11) | 21 (40) | 0 (0) |
| Thoracic surgery, n (%)<sup>c</sup> | 16 (8) | 16 (30) | 0 (0) |
| Pulmotoxic radiotherapy, n (%)<sup>d</sup> | 27 (14) | 27 (51) | 0 (0) |
| HSCT | 25 (13) | 25 (47) | 0 (0) |
| Autologous | 13 (7) | 13 (25) | 0 (0) |
| Allogeneic | 12 (6) | 12 (23) | 0 (0) |
| Asthma | 11 (6) | 2 (4) | 9 (7) |
| Bronchiolitis obliterans | 1 (<1) | 1 (2) | 0 (0) |

<sup>a</sup> BMI z-score calculated using the WHO reference population, categories defined as being underweight if z-score < -2, normal weight z-score -2 – 1, overweight z-score 1 – 2, and obese if z-score was > 2
<sup>b</sup> Other neoplasms include hepatic tumors, germ cell tumors, malignant epithelial neoplasms, malignant melanomas, unspecified malignant neoplasms, and Langerhans cell histiocytosis.
<sup>c</sup> Thoracoscopic resection (n = 2), wedge resection or metastasectomy of the lung (n = 4), open tumor resection including mediastinal or chest wall tumors (n = 7), complex cardiac surgery (n = 1), pleural procedure (n = 1), combined thoracoscopic and open tumor resection (n = 1).
<sup>d</sup> Radiation to the chest (mantle, mediastinal, or whole lung fields), abdomen (whole or any upper field), or total body irradiation.
Abbreviations: BMI body mass index; CNS, central nervous system; HSCT, hematopoietic stem cell transplantation; ICCC-3, International Classification of Childhood Cancer - Third Edition; IQR, inter quartile range.

### N_2_MBW results

Ventilation inhomogeneity in CCS assessed by N_2_MBW was overall comparable to reference values. Mean LCI was 6.27 (95%CI 6.17–6.37) compared to the reference value of 6.22, and fourteen participants (7%, 95%CI 5–12%) had LCI values above the ULN (Table 2). Their characteristics and PFT results are shown in Supplementary Table S2. S_ACIN_ x V_T_ and S_COND_ x V_T_ z-scores were slightly elevated on average, with mean z-scores of 0.35 (95%CI 0.14–0.57) and 0.10 (95%CI −0.03–0.23). N_2_MBW parameters did not differ much between high-risk and standard-risk CCS, with mean LCI z-scores of 0.22 vs −0.03, S_ACIN_ x V_T_ z-scores of 0.35 vs 0.35, and S_COND_ x V_T_ z-scores of 0.17 vs 0.07.

**Table 2.**
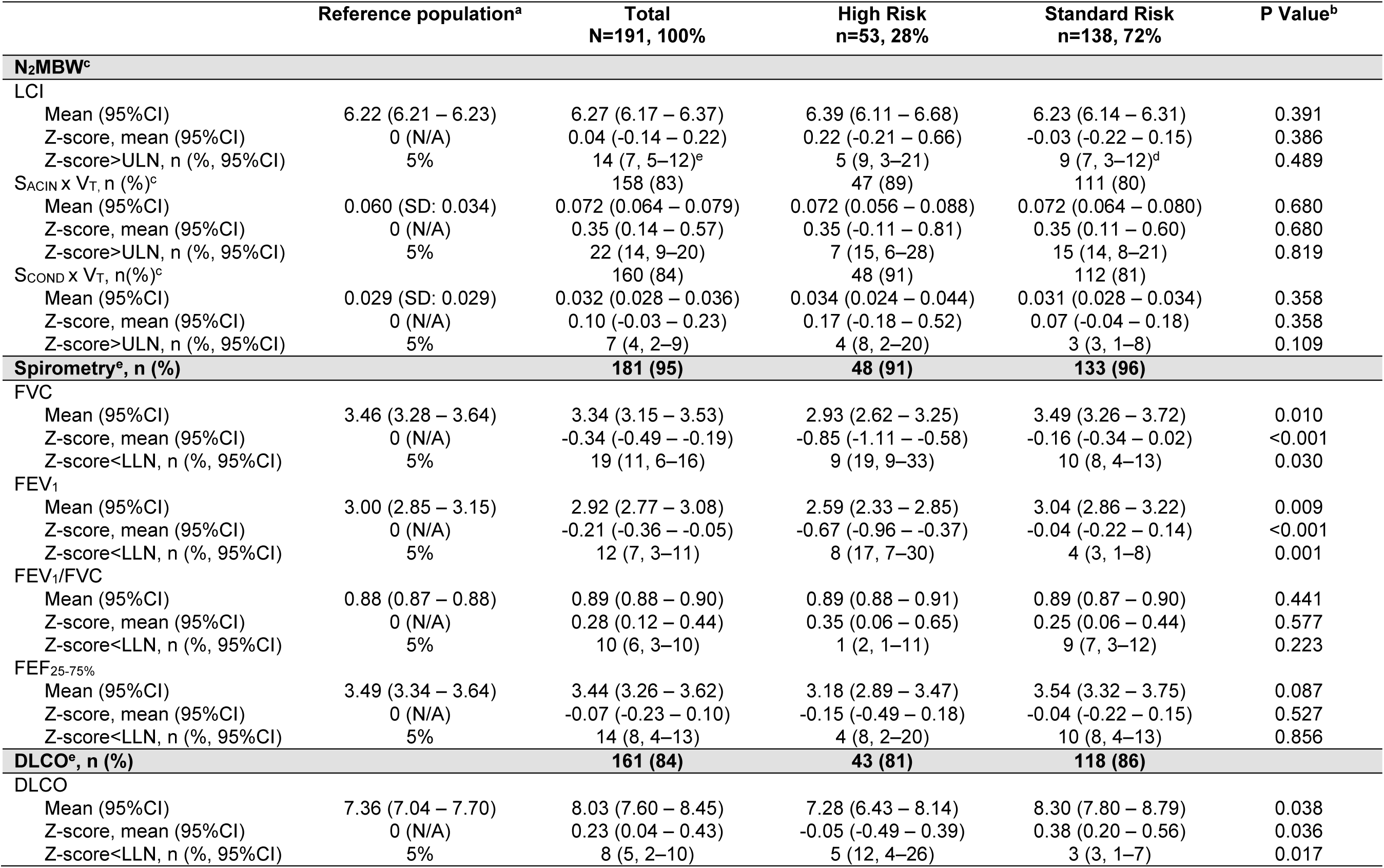
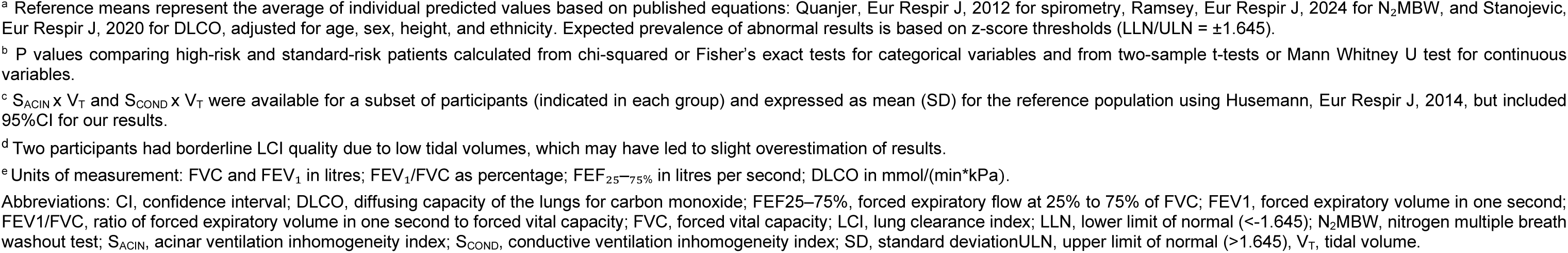
Pulmonary function parameters and impairments in childhood cancer survivors, comparing standard risk (no exposure to pulmotoxic treatments) and high-risk groups (exposed to pulmotoxic chemotherapy, radiotherapy, HSCT, and/or chest surgery)

When further stratifying the high-risk group, we found the highest mean LCI in survivors after allogeneic HSCT (7.18, 95%CI 5.99–8.38). Their mean LCI z-score was 1.58 (95%CI 0.05–3.11) and exceeded the ULN in 33% (95%CI 10–65%) (Supplementary Table S3, Figure 1). In contrast, survivors after autologous HSCT and those exposed to pulmotoxic treatments without HSCT had mean LCI values similar to the reference population (6.12 and 6.18), and few or none had abnormal results. For S_ACIN_ × V_T_, the highest mean z-score was also observed in the allogeneic HSCT group (0.99, 95% CI −0.70–2.67).

**Figure 1.**
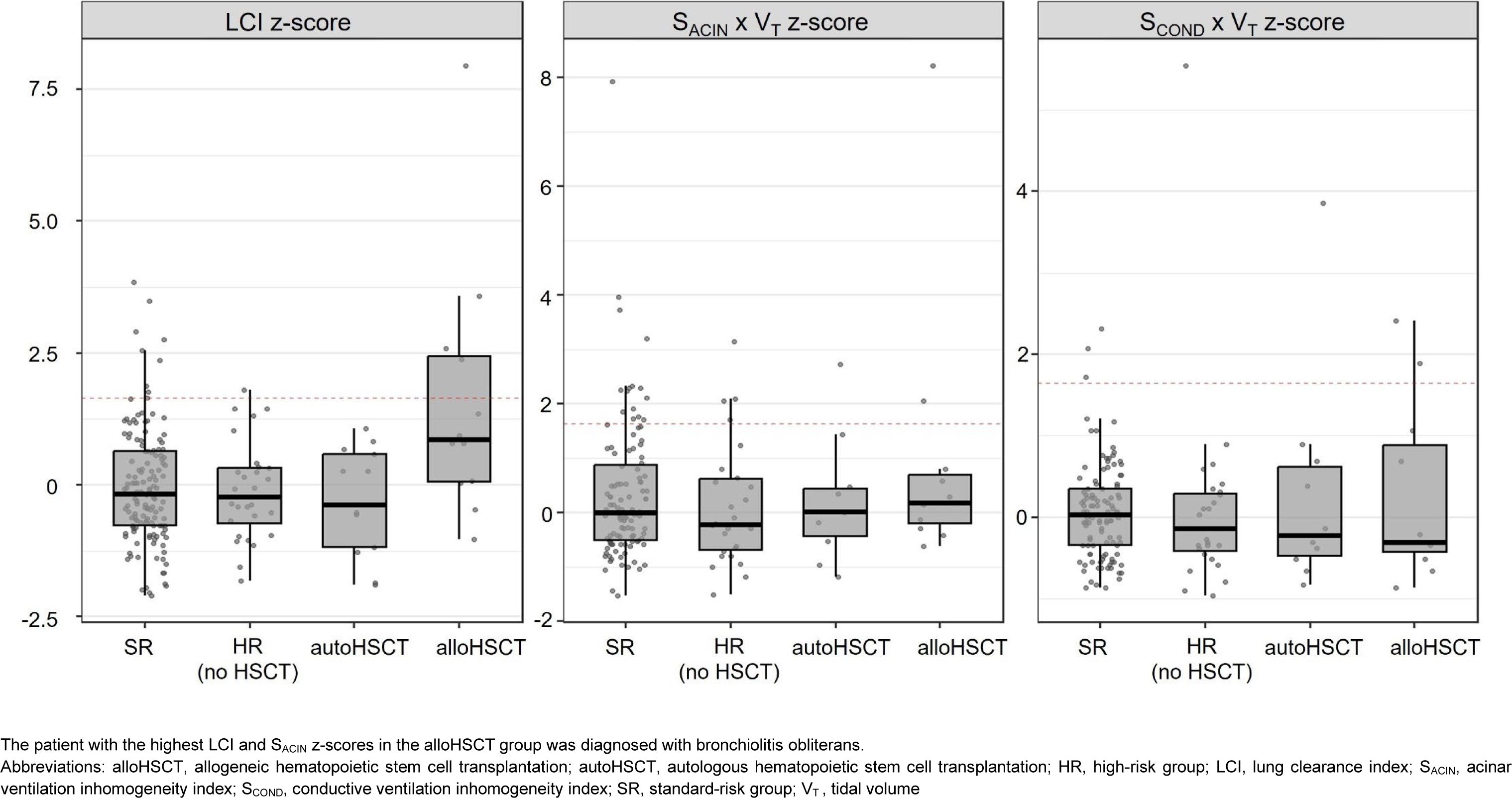
Ventilation indices (LCI, S_ACIN_ x V_T_, and S_COND_ x V_T_) in 191 childhood cancer survivors stratified into standard-risk, high-risk (no HSCT), autologous HSCT, and allogeneic HSCT groups. Dotted red lines mark the upper limit of normal (z = 1.645).

### Spirometry and DLCO results

Spirometry results of sufficient quality were available for 181 CCS. Overall, survivors had mildly decreased mean FVC and FEV_1_ z-scores of −0.34 (95%CI −0.49 – −0.19) and −0.21 (95%CI - 0.36 – −0.05), with preserved FEV_1_/FVC ratios (mean z-score 0.28) (Table 2). High-risk survivors showed more pronounced reductions in mean FVC and FEV_1_ z-scores (−0.85 and - 0.67) compared to −0.16 and −0.04 in the standard-risk group. The proportion of participants with FVC and FEV_1_ below the LLN was higher in the high-risk group (19% and 17%) than in the standard-risk group (8% and 3%; p = 0.030 and p = 0.001). FEV_1_/FVC ratios and FEF_25-75%_ were within the expected range across both groups. Obstructive patterns, defined as FEV_1_/FVC z-score < −1.645 with FVC z-score > −1.645 (28), were found in 8 participants (4%). DLCO was available in 161 (84%) participants and was lower in high-risk compared to standard-risk survivors (mean z-score −0.05 vs 0.38, p = 0.036). Nine (6%) participants had diffusion capacity impairment defined as DLCO z-score < −1.645 (28).

### LCI compared with spirometry and DLCO

LCI z-scores were inversely correlated with FEV_1_, FVC, FEV_1_/FVC, FEF_25-75%_, and DLCO, with correlation coefficients ranging from −0.16 to −0.25 (Figure 2). Pairwise scatterplots illustrate the relationship between LCI and each parameter. Among participants with elevated LCI (n=14), most did not have accompanying abnormalities in spirometry or DLCO (upper-right quadrant), varying across parameters from 8 to 11 participants. Concurrent impairment (upper-left quadrant) was less common, ranging from 2 to 5 participants. Overall, the majority of participants clustered in the lower-right quadrant, with normal LCI and normal spirometry/DLCO measures.

**Figure 2.**
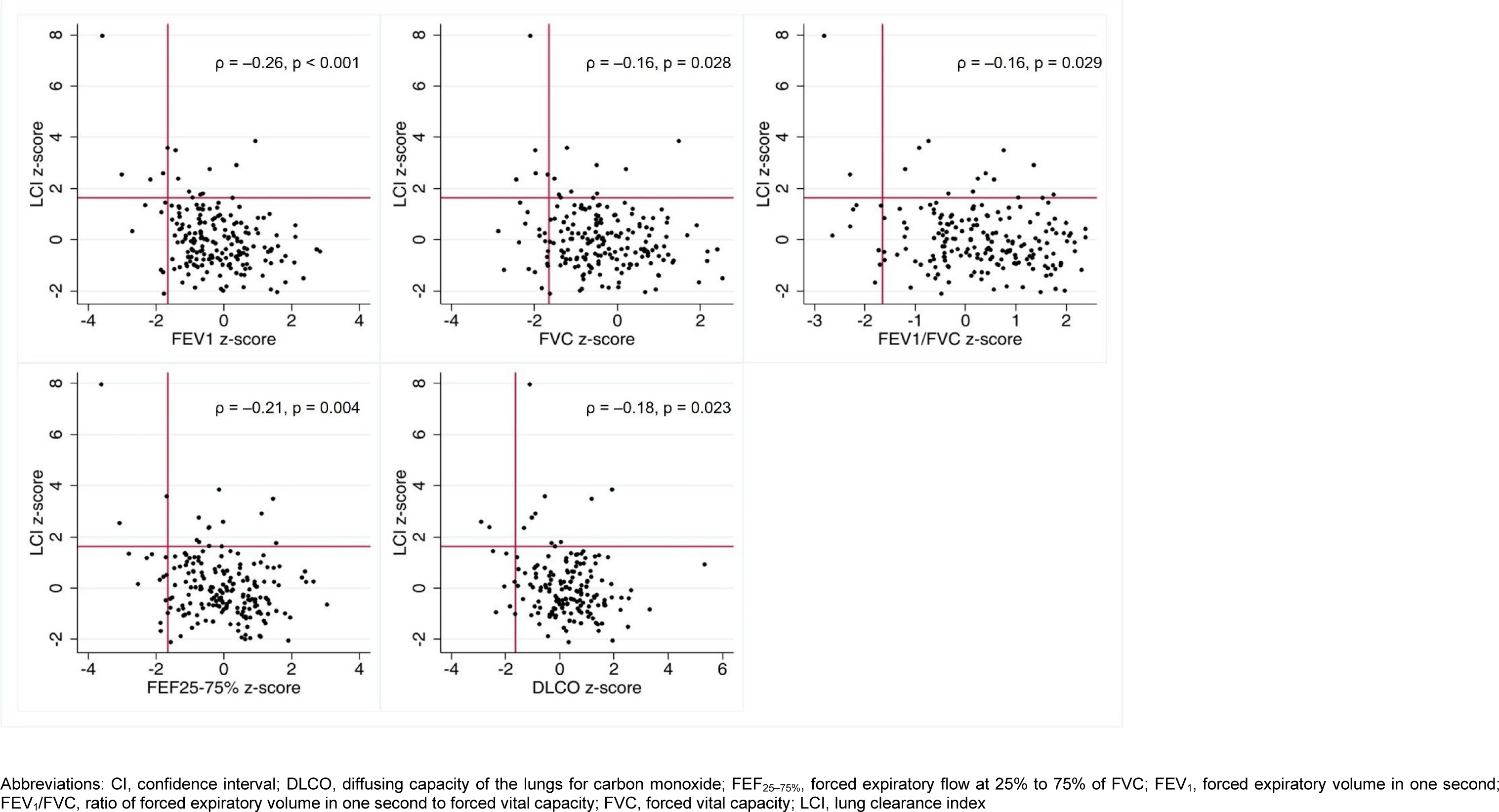
Associations between LCI z-scores and standard lung function parameters (FEV_1_, FVC, FEV_1_/FVC, FEF_25–75%_, DLCO). Scatter plots showing the relationship between LCI z-scores (y-axis) and individual lung function z-scores (x-axis). Each panel represents one comparison: FEV_1_, FVC, FEV_1_/FVC, FEF_25–75%_, and DLCO (hemoglobin-corrected). Red lines at LCI z = +1.645 and lung function z = –1.645 indicate thresholds of abnormality for respective parameters. Correlation coefficients (Spearman’s ρ) and p-values were derived using Spearman’s rank correlation test (two-sided).

In the total cohort (n=191), 36 participants had impairment in at least one spirometry or DLCO parameter, of whom 7 also had elevated LCI. An additional 7 participants showed isolated LCI impairment without other abnormalities; however, in 2 of these, N_2_MBW test quality was borderline due to low tidal volumes. Thus, only 5 participants (3% of the entire cohort) had isolated and technically reliable LCI impairment.

### Associations between treatment exposures and N₂MBW parameters

In multivariable analysis, allogeneic HSCT was associated with higher LCI z-scores (1.42, 95%CI 0.57 to 2.26) and increased acinar ventilation inhomogeneity (S_ACIN_ × V_T_ z-score: 1.27, 95%CI 0.25 to 2.29) (Table 3). We found no associations with other treatment exposures, including autologous HSCT, pulmotoxic chemotherapy, thoracic surgery, or pulmotoxic radiotherapy. Asthma was associated with elevated LCI (1.21, 95%CI 0.46 to 1.96) and S_ACIN_ × V_T_ z-scores (1.60, 95%CI 0.62 to 2.52).

**Table 3.**
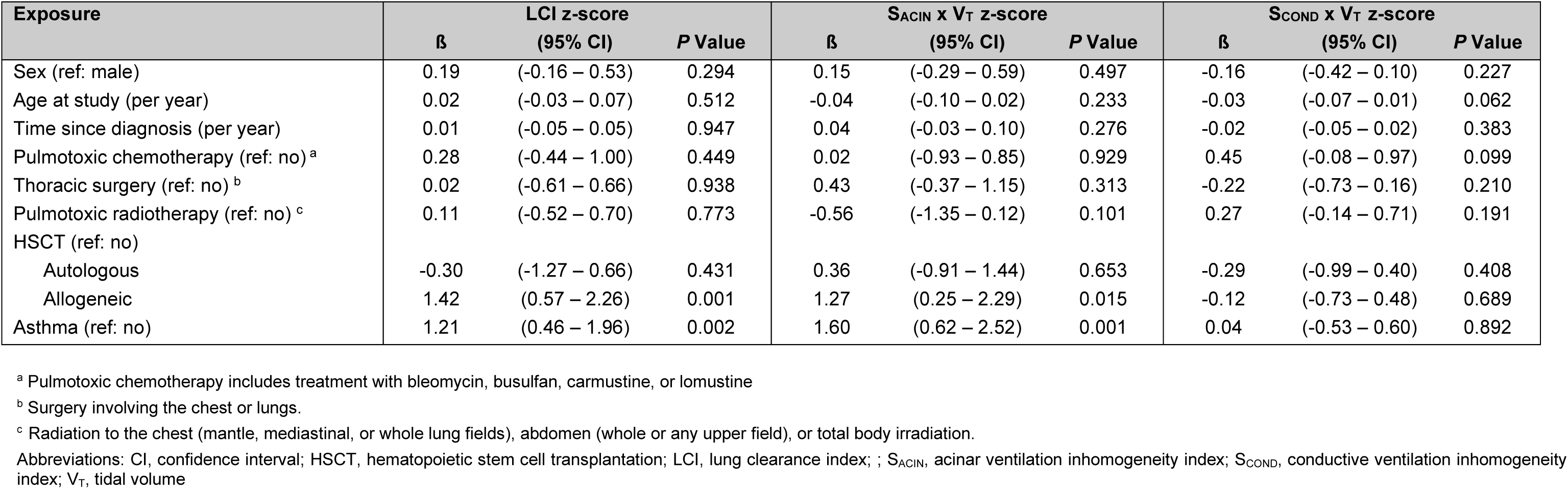
Clinical and treatment predictors of lung clearance and ventilation inhomogeneity in childhood cancer survivors. Multivariable linear regression results for LCI, S_ACIN_, and S_COND_ (z-scores).

| Exposure | LCI z-score |  |  | S<sub>ACIN</sub> x V<sub>T</sub> z-score |  |  | S<sub>COND</sub> x V<sub>T</sub> z-score |  |  |
| --- | --- | --- | --- | --- | --- | --- | --- | --- | --- |
| | $\beta$ | (95% CI) | P Value | $\beta$ | (95% CI) | P Value | $\beta$ | (95% CI) | P Value |
| Sex (ref: male) | 0.19 | (-0.16 – 0.53) | 0.294 | 0.15 | (-0.29 – 0.59) | 0.497 | -0.16 | (-0.42 – 0.10) | 0.227 |
| Age at study (per year) | 0.02 | (-0.03 – 0.07) | 0.512 | -0.04 | (-0.10 – 0.02) | 0.233 | -0.03 | (-0.07 – 0.01) | 0.062 |
| Time since diagnosis (per year) | 0.01 | (-0.05 – 0.05) | 0.947 | 0.04 | (-0.03 – 0.10) | 0.276 | -0.02 | (-0.05 – 0.02) | 0.383 |
| Pulmonary chemotherapy (ref: no) <sup>a</sup> | 0.28 | (-0.44 – 1.00) | 0.449 | 0.02 | (-0.93 – 0.85) | 0.929 | 0.45 | (-0.08 – 0.97) | 0.099 |
| Thoracic surgery (ref: no) <sup>b</sup> | 0.02 | (-0.61 – 0.66) | 0.938 | 0.43 | (-0.37 – 1.15) | 0.313 | -0.22 | (-0.73 – 0.16) | 0.210 |
| Pulmonary radiotherapy (ref: no) <sup>c</sup> | 0.11 | (-0.52 – 0.70) | 0.773 | -0.56 | (-1.35 – 0.12) | 0.101 | 0.27 | (-0.14 – 0.71) | 0.191 |
| HSCT (ref: no) |  |  |  |  |  |  |  |  |  |
| Autologous | -0.30 | (-1.27 – 0.66) | 0.431 | 0.36 | (-0.91 – 1.44) | 0.653 | -0.29 | (-0.99 – 0.40) | 0.408 |
| Allogeneic | 1.42 | (0.57 – 2.26) | 0.001 | 1.27 | (0.25 – 2.29) | 0.015 | -0.12 | (-0.73 – 0.48) | 0.689 |
| Asthma (ref: no) | 1.21 | (0.46 – 1.96) | 0.002 | 1.60 | (0.62 – 2.52) | 0.001 | 0.04 | (-0.53 – 0.60) | 0.892 |

<sup>a</sup> Pulmonary chemotherapy includes treatment with bleomycin, busulfan, carmustine, or lomustine
<sup>b</sup> Surgery involving the chest or lungs.
<sup>c</sup> Radiation to the chest (mantle, mediastinal, or whole lung fields), abdomen (whole or any upper field), or total body irradiation.
Abbreviations: CI, confidence interval; HSCT, hematopoietic stem cell transplantation; LCI, lung clearance index; S<sub>ACIN</sub>, acinar ventilation inhomogeneity index; S<sub>COND</sub>, conductive ventilation inhomogeneity index; V<sub>T</sub>, tidal volume

## Discussion

This is the largest prospective study to date to assess N_2_MBW in a comprehensive cohort of children and adolescents after cancer treatment, alongside spirometry and DLCO. We found that most survivors had normal lung function across all measures. Isolated LCI abnormalities were rare, correlations between LCI and spirometry or DLCO were weak, and overall, N_2_MBW did not reveal substantially more abnormalities than standard tests. However, survivors of allogeneic HSCT showed higher LCI and S_ACIN_ values, and this treatment was the only exposure significantly associated with N_2_MBW abnormalities in multivariable analysis.

The overall LCI z-score of 0.04 in our cohort indicates largely normal findings. Of the studies identified in our systematic review, only two had assessed N₂MBW in cohorts of CCS with diverse treatment exposures (12). An Italian study of 57 pediatric CCS, evaluated a median of 6 years after chemotherapy or radiotherapy, reported comparable results, with a mean LCI z-score of 0.002 (29). In contrast, our previous study of adult Swiss CCS, who were on average 20 years from diagnosis, found substantially higher LCI values in both high- and standard-risk groups defined as in the present study (13). In that adult cohort, mean LCI z-scores were 2.09 and 0.95 in the high- and standard-risk groups, compared with 0.26 and −0.05 in the present study. The prevalence of abnormal LCI was also markedly higher in both risk groups of adults (60% and 23%) than in our pediatric cohort (9% and 7%). Since the publication of the systematic review, one additional study in Danish survivors of acute lymphoblastic leukemia (n = 146, mean 7 years post-diagnosis) reported slightly higher mean LCI values of 6.71 (z-score 0.91) (30). Comparisons across studies should be made cautiously, as each used different reference equations for z-scores, which may partly explain discrepancies. The available data suggest that, in younger survivors, lung injury may still be below the threshold of detection by N_2_MBW. In contrast, the greater impairments observed in adult survivors could indicate that ventilation inhomogeneity emerges or worsens over time, a possibility supported by the Italian study’s positive correlation between LCI and time since diagnosis (29). Alternatively, these differences may reflect treatment-era effects, with earlier regimens being more intensive or toxic and leading to more pronounced long-term lung damage.

When stratifying our cohort by treatment exposures, ventilation inhomogeneity indices (LCI, S_ACIN_, and S_COND_) were largely comparable between the standard-risk group, the high-risk group with exposures other than HSCT, and the autologous HSCT recipients. However, survivors of allogeneic HSCT had a substantially increased prevalence of abnormal LCI values (33%, 95%CI 10–65%), exceeding the expected 5% in a healthy reference population. This aligns with previous studies in pediatric survivors of allogeneic HSCT, after various underlying diagnoses, which have reported increased LCI even among asymptomatic survivors with no spirometric abnormalities (31–35). The pulmonary sequelae in this population are often driven by chronic graft-versus-host disease and associated inflammatory processes, which can lead to small airway fibrosis and bronchiolitis obliterans, characterized by obstruction and ventilation inhomogeneity, particularly affecting distal airways. Although N_2_MBW may be sensitive to these subtle changes, impairments in conventional measures such as FEV_1_ and DLCO were also common in allogeneic HSCT survivors (30% and 27%, respectively), with no cases of isolated LCI abnormalities. This overlap suggests that, in this cohort, N_2_MBW did not detect additional impairment beyond standard lung function tests. Nonetheless, its potential to capture small airway involvement or to identify changes before declines in spirometry or DLCO become evident remains to be further assessed in larger samples and longitudinal studies.

Also, the multivariable linear regression suggested that only allogeneic HSCT was associated with increased ventilation inhomogeneity, and no other lung-damaging treatments. This may reflect the limited subgroup sizes and the relatively short median interval of seven years since diagnosis, which may be too early for N_2_MBW-detectable ventilation abnormalities to develop. Pulmonary effects of radiotherapy and certain chemotherapies often evolve gradually and may become apparent only years or decades after treatment (36,37). Larger cohorts and longitudinal studies will be essential to clarify the delayed impact of these treatments. We also observed elevated LCI and S_ACIN_ in participants with asthma, which reflects small airway involvement and patchy airway narrowing characteristic of this condition and is consistent with previous reports (38,39). Among the 14 participants with elevated LCI, 5 (36%) had asthma, suggesting that asthma may contribute substantially to ventilation inhomogeneity even among CCS. However, as tests such as FeNO, bronchodilator response, or bronchial challenge were not systematically performed, we cannot be certain of the underlying cause of the abnormalities. Future studies and clinical assessments should therefore account for asthma as a potential contributor to impairment and include measures to distinguish it from treatment-induced lung injury.

The modest correlations between N_2_MBW parameters and standard lung function tests suggest these tests capture overlapping yet distinct aspects of pulmonary pathology. LCI was only weakly correlated with spirometric indices and diffusion capacity, while isolated ventilation inhomogeneity was uncommon. This suggests that small airway dysfunction detectable by N_2_MBW is rare in this cohort and usually accompanies other lung function impairments. Standard tests primarily revealed restrictive and diffusion abnormalities. These findings likely reflect underlying pathophysiological processes such as interstitial changes or microvascular injury, which reduce gas transfer and lung volumes without significantly affecting peripheral airway patency or causing ventilation inhomogeneity. Interstitial lung injury is often the final common pathway of various lung-damaging cancer treatments and can present as chronic interstitial pneumonia that may progress to lung fibrosis (40). This aligns with previous evidence indicating that diffusion and restrictive abnormalities predominate in CCS, as long-term studies have shown substantially higher rates of restriction (18–42%) and impaired DLCO (34–40%) in high-risk CCS, while obstructive patterns remained uncommon (2–8%) (41–44). N_2_MBW may therefore fail to capture such early interstitial fibrotic changes or vascular damage, especially in younger survivors shortly post-treatment.

This study is strengthened by a high participation rate, limiting selection bias. PFTs were performed under standardized conditions in specialized laboratories and underwent rigorous quality control. Technicians were blinded to treatment groups, and detailed treatment data allowed us to explore associations with N_2_MBW outcomes. The inclusion of a well-characterized pediatric cohort shortly after treatment contributes valuable insight into pulmonary function in the early phase of survivorship. However, including only one measurement timepoint limits understanding of the temporal development of lung impairment, and variable follow-up durations may have introduced heterogeneity. Interpretation of S_ACIN_ and S_COND_ results is complicated by their known sensitivity to breathing pattern variability and the absence of robust pediatric reference values. In this study, we used reference values derived from a small cohort of healthy young adults (22), which may limit their generalizability to a pediatric population. Subgroups were small, resulting in wide confidence intervals and limited statistical power to detect subtle associations.

In summary, this large pediatric CCS cohort, seven years post-diagnosis, suggests that N_2_MBW may provide limited additional information beyond standard lung function tests. Ventilation inhomogeneity was uncommon and largely confined to survivors of allogeneic HSCT, where N_2_MBW may offer supplementary insight into early small airway changes. Given the largely normal findings in other survivors, our results do not currently support the routine incorporation of N_2_MBW into standard clinical follow-up. However, prior evidence from adult CCS suggests that N_2_MBW abnormalities may emerge or progress over time, even in CCS not treated with allogeneic HSCT. Therefore, we need longitudinal studies with larger, well-characterized cohorts to clarify the timing, trajectory, and prognostic significance of N_2_MBW abnormalities, and their potential role in guiding risk-adapted follow-up.

## Data Availability

All data produced in the present study are available upon reasonable request to the authors.

## Statements & Declarations

## Acknowledgements

We thank all survivors for participating in our study. We thank the Pediatric Hematology/Oncology and Pediatric Pulmonology teams at the University Children’s Hospitals in Bern, Basel, and Geneva, especially study nurses Sandra Lüscher, Artemis Ionnaki, Valentine Pradet, and Andrea Ziörjen, and the study team of the Childhood Cancer Research Group.

This paper incorporates suggestions from AI tools, including Grammarly and ChatGPT (OpenAI), which were used to improve the clarity, grammar, and readability of the written text. All scientific content, interpretation, and conclusions are the responsibility of the authors.

## Author contribution

MŽ: formal analysis, writing—original draft preparation, visualization; CS: writing—review and editing; NW: writing—review and editing; JU: writing—review and editing, CS: writing—review and editing; AM: writing—review and editing, MA: writing—review and editing; PL: writing—review and editing; CEK: conceptualization, methodology, writing— review and editing, funding acquisition, supervision.

## Ethics approval

The Ethics Committee of the Canton of Bern (2019–00739) granted ethical approval.

---

**Supplementary Figure S1.**
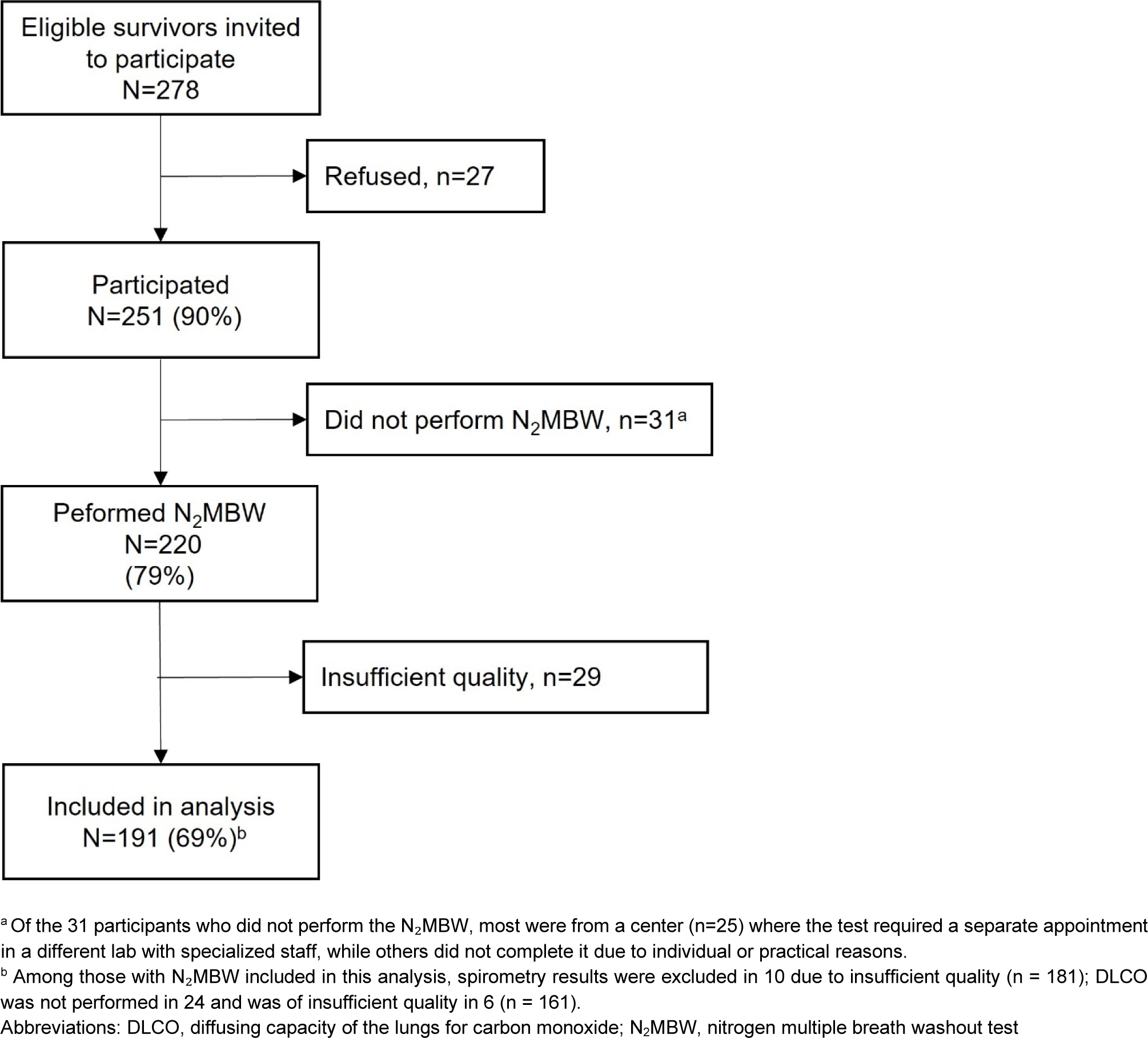
Flow diagram of the study population starting with survivors who were eligible and invited to participate.

**Supplementary Table S1.** Demographic and clinical characteristics of childhood cancer survivors in the SCCSS FollowUp-Pulmo study, stratified by availability of nitrogen multiple-breath washout assessment.

|  | Total SCCSS<br>FollowUp-Pulmo<br>N=251 | N<sub>2</sub>MBW<br>assessment<br>N=191 | No N<sub>2</sub>MBW<br>assessment<br>n=60 | P Value<sup>a</sup> |
| --- | --- | --- | --- | --- |
| **Demographic characteristics** |  |  |  |  |
| Sex |  |  |  | 0.987 |
| Male, n (%) | 142 (57) | 108 (57) | 34 (57) |  |
| Female, n (%) | 109 (43) | 83 (43) | 26 (43) |  |
| Age at study in years, median (IQR) | 14 (10–17) | 14 (11–17) | 13 (9–16) | 0.021 |
| Age at diagnosis in years, median (IQR) | 5 (3–10) | 5 (3–10) | 5 (3–10) | 0.792 |
| Time since diagnosis in years, median (IQR) | 7 (4–10) | 7 (5–10) | 5 (3–9) | 0.036 |
| **Clinical characteristics** |  |  |  |  |
| ICCC-3 cancer diagnosis, n (%) |  |  |  | 0.536 |
| Leukemia | 124 (49) | 91 (48) | 33 (55) |  |
| Lymphoma | 30 (12) | 24 (13) | 6 (10) |  |
| CNS neoplasms | 14 (6) | 9 (5) | 5 (8) |  |
| Neuroblastoma | 22 (9) | 20 (11) | 2 (3) |  |
| Renal tumor | 20 (8) | 16 (8) | 4 (7) |  |
| Malignant bone tumor | 10 (4) | 7 (4) | 3 (5) |  |
| Soft tissue sarcomas | 16 (6) | 12 (6) | 4 (7) |  |
| Other neoplasms<sup>b</sup> | 15 (6) | 12 (6) | 3 (5) |  |
| History of relapse, n (%) | 21 (9) | 15 (8) | 6 (10) | 0.575 |
| Pulmotoxic chemotherapy, n (%)<sup>c</sup> | 28 (11) | 21 (11) | 7 (12) | 0.885 |
| Thoracic surgery, n (%) | 23 (9) | 20 (11) | 3 (5) | 0.200 |
| Pulmotoxic radiotherapy, n (%)<sup>d</sup> | 35 (14) | 27 (14) | 8 (13) | 0.876 |
| HSCT | 33 (14) | 25 (13) | 8 (13) | 0.961 |
| Autologous | 15 (6) | 13 (7) | 2 (4) |  |
| Allogeneic | 18 (7) | 12 (6) | 6 (10) |  |
| Asthma | 16 (6) | 11 (6) | 5 (8) | 0.476 |

<sup>a</sup> P values comparing high-risk and standard-risk patients calculated from chi-squared or Fisher's exact tests for categorical variables and from t-tests for continuous variables.
<sup>b</sup> Other neoplasms contain hepatic tumors, germ cell tumors, malignant epithelial neoplasms, malignant melanomas, unspecified malignant neoplasms, and Langerhans cell histiocytosis.
<sup>c</sup> Pulmotoxic chemotherapy includes treatment with bleomycin, busulfan, carmustine, or lomustine
<sup>d</sup> Pulmotoxic radiotherapy includes radiation to the thorax, upper abdomen, and total body irradiation
Abbreviations: BMI body mass index; CNS, central nervous system; HSCT, hematopoietic stem cell transplantation; ICC-3, International Classification of Childhood Cancer - Third Edition; IQR, inter quartile range.

**Supplementary Table S2.** Characteristics of childhood cancer survivors with abnormal lung clearance index (z-score > 1.645), including treatment exposures and pulmonary function parameters.

|  | Sex | Age group | Childhood cancer diagnosis | HSCT | Pulmotoxic chemotherapy | Pulmotoxic radiotherapy | LCI | LCI z-score | S<sub>ACIN</sub> X V<sub>T</sub> z-score | S<sub>COND</sub> x V<sub>T</sub> z-score | FEV<sub>1</sub> z-score | FVC z-score | DLCO z-score | Pulmonary comorbidity |
| --- | --- | --- | --- | --- | --- | --- | --- | --- | --- | --- | --- | --- | --- | --- |
| 1 | M | 15-20 | Leukemia | - | - | - | 7.28 | 2.54 | - | - | -3.01 | -1.68 | - | Asthma |
| 2 | M | 15-20 | Leukemia | - | - | - | 7.81 | 3.49 | 3.97 | -0.10 | -1.42 | -1.98 | 1.18 | Asthma |
| 3<sup>a</sup> | F | 10-15 | Leukemia | - | - | - | 7.04 | 1.76 | 2.12 | 0.76 | -0.69 | -1.41 | -0.31 | - |
| 4 | F | 5-10 | Neuroblastoma | - | - | - | 7.48 | 1.88 | -0.29 | 0.62 | -1.02 | -1.11 | - | Asthma |
| 5 | F | 15-20 | Soft tissue sarcoma | - | - | - | 7.24 | 2.35 | - | - | -2.17 | -2.44 | -1.32 | - |
| 6 | M | 10-15 | Leukemia | Allogeneic | - | TBI | 7.47 | 2.39 | -0.12 | 1.90 | -1.34 | -1.52 | -2.60 | - |
| 7 | F | 15-20 | CNS tumor | - | - | - | 6.92 | 1.65 | 2.24 | -0.55 | -0.92 | -1.37 | - | - |
| 8 | M | 10-15 | Leukemia | - | - | - | 8.16 | 3.85 | 1.44 | 2.31 | 0.93 | 1.48 | 1.93 | Asthma |
| 9<sup>a</sup> | M | 15-20 | Leukemia | - | - | - | 7.47 | 2.92 | 1.74 | 2.07 | 0.38 | -0.50 | -0.90 | - |
| 10 | F | 15-20 | Leukemia | Allogeneic | - | TBI | 7.94 | 3.59 | 0.15 | 2.41 | -1.66 | -1.22 | -0.55 | - |
| 11 | M | 15-20 | Lymphoma | - | - | Chest | 6.92 | 1.80 | 2.09 | -0.79 | -0.62 | -0.49 | 0.04 | Asthma |
| 12 | F | 5-10 | Leukemia | Allogeneic | Busulfan | - | 12.78 | 7.96 | 8.24 | 0.69 | -3.58 | -2.1 | -1.11 | Bronchiolitis obliterans |
| 13 | F | 10-15 | Leukemia | Allogeneic | - | TBI | 7.44 | 2.60 | 2.06 | -0.66 | -1.79 | -1.97 | -2.90 | - |
| 14 | F | 15-20 | Leukemia | - | - | - | 7.56 | 2.76 | -0.29 | -0.66 | -0.42 | 0.21 | -1.03 | - |

<sup>a</sup> Participants with borderline LCI quality due to low tidal volumes, which may have led to slight overestimation of results.
Abbreviations: CNS, central nervous system; DLCO, diffusing capacity of the lungs for carbon monoxide; F, female; FEV<sub>1</sub>, forced expiratory volume in 1 second; FVC, forced vital capacity; HSCT, hematopoietic stem cell transplantation; LCI, lung clearance index; M, male; S<sub>ACIN</sub>, acinar ventilation inhomogeneity index; S<sub>COND</sub>, conductive ventilation inhomogeneity index; TBI, total body irradiation

**Supplementary Table S3.**
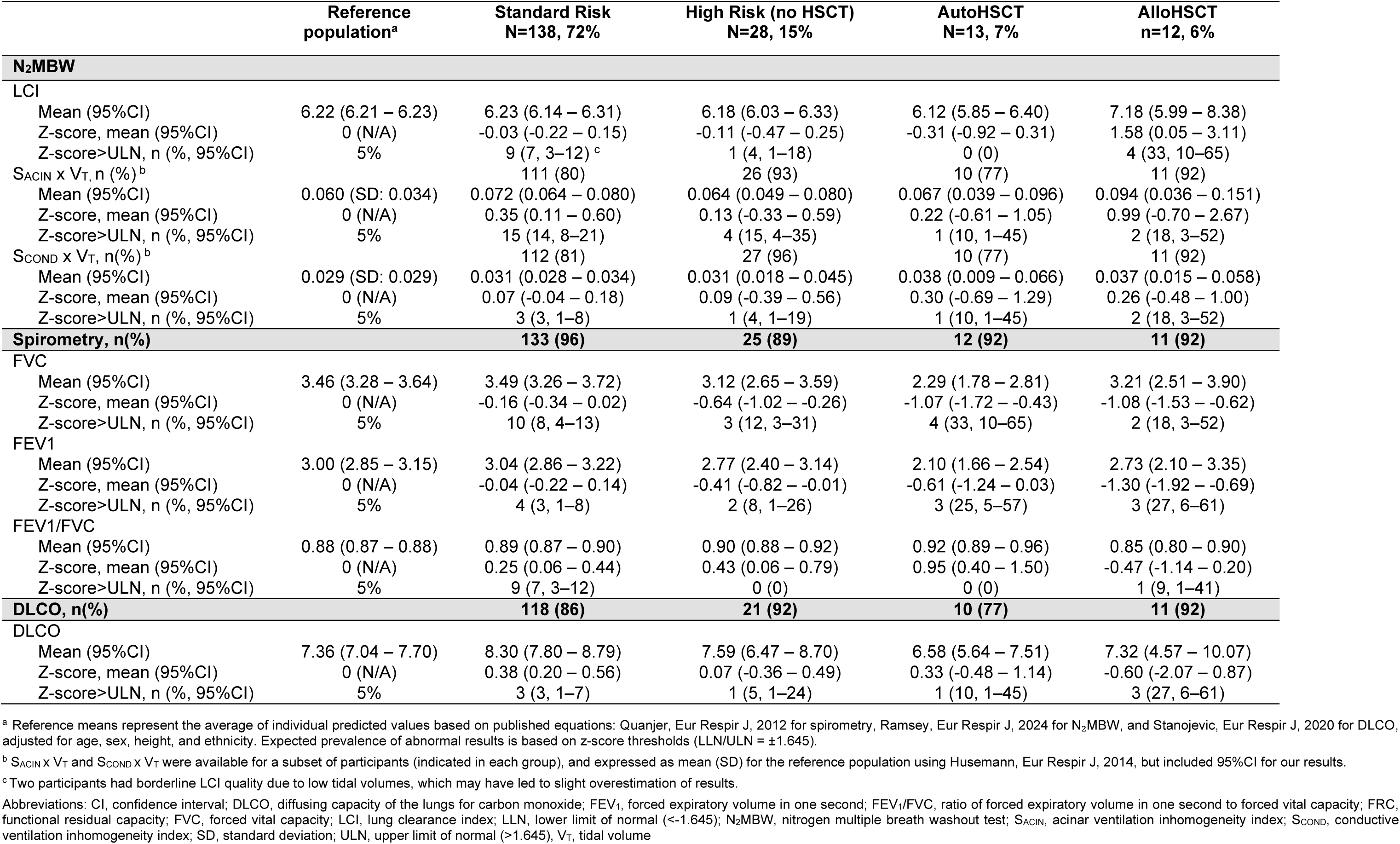
Comparison of nitrogen multiple-breath washout indices between standard-risk childhood cancer survivors, high-risk without HSCT, high-risk who underwent autologous HSCT, and high-risk who underwent allogeneic HSCT. (N = 191)

